# Community Perceptions, Knowledge, and Experiences of Pre-eclampsia: An Exploratory Study in Uganda

**DOI:** 10.1101/2024.11.30.24317454

**Authors:** Enid Kawala Kagoya, Allan G Nsubuga, Irene Authairwe, Prossy Nakatudde, Catherine Asiimwe, Chispus Gidudu, Elizabeth Ajalo, Paul Waako, Julius Wandabwa, Lawrence Arach, Grace Mbabazi Atwakire, Milton Musaba, Ronald Kibuuka, Faith Nyangoma, Sheilla Mbanago, Joshua Mugabi, Violet Chemutai, Jesca Atugonza, Byron Jonathan Ewaala, Betty Nakawuka, Francis Okello, Richard Mugahi, Akello Jackline, Andrew Twineamatsiko, Moses Adroma, Kenneth Mugabe

## Abstract

**Background:** Pre-eclampsia and related hypertensive disorders of pregnancy are the second leading cause of maternal mortality globally, with 95% of the burden in low and middle in countries. In Uganda, these conditions account for 16% of maternal deaths. Pre-eclampsia, which affects 2-8% of pregnancies worldwide, significantly impacts both maternal and perinatal health due to complications and iatrogenic preterm delivery. Effective management hinges on understanding the condition, early symptom recognition, and timely healthcare seeking. However, patient perspectives and experiences on pre-eclampsia in low-and middle-income countries remain underexplored.

**Objective:** This study aimed to explore knowledge, myths, and experiences related to pre-eclampsia among women of reproductive age in Mbale city, Eastern Uganda, to understand their impact on health-seeking behaviours.

**Methods:** An explorative qualitative study was conducted using in-depth interviews with 81 women aged 18-49 years, recruited during a community outreach initiative on pre-eclampsia. Data were collected over 6 days between 21^st^-26^th^/May/2024 using a semi-structured interview guide. Interviews were transcribed and analyzed thematically using ATLAS.ti software.

**Results:** Participants demonstrated a limited understanding of pre-eclampsia, with common misconceptions including associating it with swollen feet, body weakness, marital distress, multiple pregnancies, high blood pressure and witchcraft reflecting a mix of accurate and misguided understandings. Participants showed varying responses to pre-eclampsia, with some relying on traditional remedies and others seeking modern medical care. Management strategies included herbal treatments and unconventional remedies.

**Conclusion:** The study highlights a critical gap in accurate knowledge and awareness of pre-eclampsia among women in Mbale City. Misconceptions and reliance on traditional medicine contribute to delays in seeking appropriate care. Culturally tailored educational interventions are needed to improve awareness, and understanding of pre-eclampsia, and promote timely medical care, essential for improving maternal and neonatal outcomes in low-income settings.

## Background

Preeclampsia, eclampsia and other hypertensive disorders of pregnancy are the second leading cause of maternal mortality worldwide[1]. This burden is most pronounced in low- and middle-income countries which account for nearly 95% of maternal deaths[2]. Sub-Saharan Africa alone accounts for 70% of these deaths, with nearly 25% attributed to hypertensive disorders of pregnancy[2]. In Uganda, hypertensive disorders are the second leading cause of maternal mortality, contributing to 16% of maternal deaths [3]. Pre-eclampsia, which complicates 2-8% of pregnancies globally, is the main cause of mortality and morbidity[4]. It also leads to significant perinatal mortality and morbidity due to increased risk of iatrogenic preterm delivery, as delivery of the placenta is considered the definitive cure of pre-eclampsia[3]. Effective management through early symptom recognition, timely presentation to health facilities, antihypertensive treatment, magnesium sulphate administration, and prompt delivery can improve maternal and newborn outcomes of pregnancies complicated by pre-eclampsia[4]. Early symptom recognition and timely health facility presentation are critically influenced by patients’ knowledge, perceptions and experiences[5]. Studies show that a higher level of knowledge about pre-eclampsia correlates with an increased likelihood of seeking appropriate care [6]. Despite the importance of knowledge, there is limited research on patient perspectives regarding pre-eclampsia, particularly in low-and middle-income settings [2]. Women’s knowledge and experiences with pre-eclampsia in these settings remain largely unknown. This study aimed to address this gap by exploring the myths, knowledge and experiences related to pre-eclampsia and eclampsia among women of reproductive age in Mbale City, Eastern Uganda. A better understanding of women’s knowledge and experiences with pre-eclampsia and eclampsia is crucial for improving health-seeking behaviors and outcomes for high-risk populations [5].

## Materials and methods

### Study design

This qualitative study followed an exploratory narrative design, using in-depth interviews.

### Study site

This study was conducted in Mbale City, a newly approved city in southeastern Uganda. Mbale is located at the western foot of Mount Elgon (4,321 meters), 120 kilometers northeast of Jinja. Known for its fertile coffee-growing region, Mbale serves as an agricultural trade Center and is home to one of Uganda’s principal dairies and regional referral hospitals.

### Study population

We initially aimed to interview women of reproductive age. The final study population consisted of women aged 18 to 49 years.

### Data collection

The study participants were recruited during a community outreach initiative aimed at raising awareness about pre-eclampsia in Mbale City, which included blood pressure measurements for attendees. Data collection occurred over 6 days between 21^st^-26^th^/May/2024, using in-depth interviews, guided by a semi-structured interview guide. The guide contained questions designed to explore women’s knowledge, understanding, perceptions and experiences regarding pre-eclampsia. Interviews were recorded and supplemented with simultaneous note-taking.

### Data management

All the interview recordings were transcribed by the study team, who read the transcripts and listened to the audio recordings multiple times to ensure consistency. The audio recorders were securely stored under a key and lock, only accessible by the research team.

### Data analysis

The transcripts were analyzed using ATLAS.ti software, employing a thematic analysis following a deductive approach. Priori coding was conducted, and the transcripts were subsequently exported into the software to aid the organization of nodes, themes and subthemes. After the vigorous process of analysis, five main themes emerged, each with their respective subthemes as detailed in the results section.

### Ethical considerations

Ethical approval was obtained from the Research Ethics Committees (REC) of Mbale Regional Referral Hospital (approval number MRRH-2023-300. Administrative permission to collect data was granted by the district health office and parish authorities gave administrative permission to collect data. Written informed consent was obtained from all participants before their inclusion in the study.

## Results

We recruited 81 participants, the majority of whom (51/81; 63%) were below the age of 35 years. Most participants (51/81; 63%) were either married or cohabiting. Nearly three-quarters of the participants (50/81, 61.7%) had heard about pre-eclampsia.

The study identified four thematic areas: knowledge of signs and symptoms, management, prevention and perceptions about preeclampsia. These themes are summarized in Figure 1 below.

**Figure 1:**
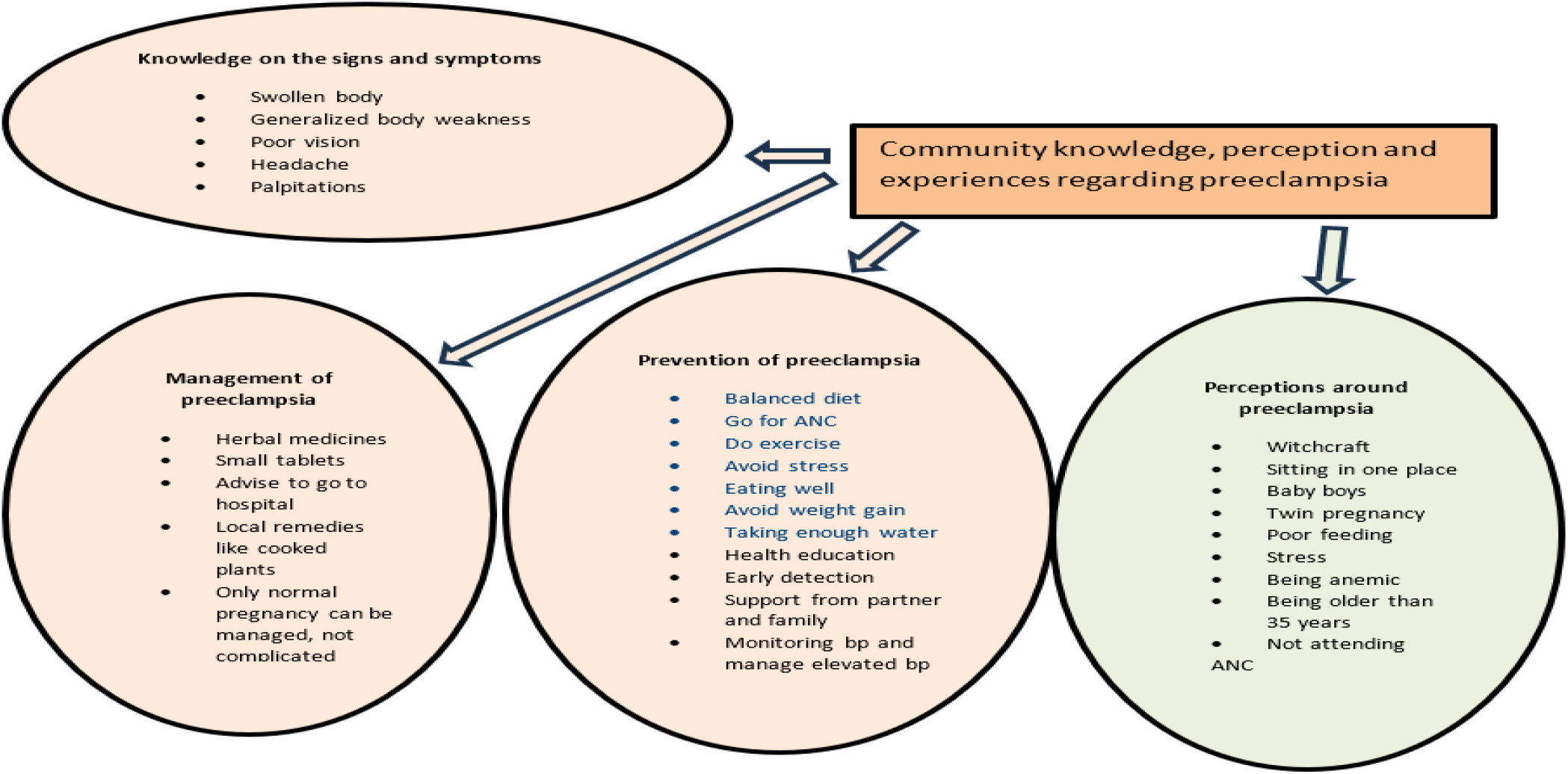
Themes identified from the transcripts.

### Theme one: Knowledge of preeclampsia, its signs and symptoms

The study revealed that most mothers had a limited understanding of pre-eclampsia and its true nature. Instead, they associated the condition primarily with observable signs and symptoms experienced during pregnancy. While many participants incorrectly believed that pre-eclampsia was related to high blood pressure resulting from stress, they accurately identified multiple pregnancies and foetal macrosomia as risk factors.

For example, one participant remarked:

> *“I think it is caused by (high) pressure, stress, and a new experience of the body,” (Mother, 15-19 years)”*

Another participant stated:

> *“It happens to those who give birth to big babies or twins; there is too much weight on the lower limbs” (Mother, 20-24 years)*

Most mothers associated pre-eclampsia with oedema, specifically “swollen feet,” which was historically considered a symptom of the condition. However, oedema is no longer a diagnostic criterion for pre-eclampsia, as it is commonly observed in normotensive pregnant women.

> *“It’s related to having swollen feet”, (Mother, 20-24 years)”*.

Additionally, one participant described pre-eclampsia as:

> *“It’s a high blood pressure that comes automatically during pregnancy”(Mother, 25-29)*.

### Subtheme one: Knowledge of susceptibility and risk factors

The study revealed that pre-eclampsia was perceived as a condition that affects all mothers, irrespective of socioeconomic status. Many participants believed that all pregnant women are at risk of preeclampsia.

> *“Not selective, whether fat or thin, young or old, everyone can get pressure” (Mother, 25-29 years)*.

Some participants were aware that mothers over 35 years of age were at a higher risk of pre-eclampsia. However, others were unaware of these risk factors, often due to not attending antenatal care during pregnancy.

> *“Pregnant mothers especially those 35 years old and above” (Mother,30-35 years)*.

Several mothers reported a lack of knowledge about pre-eclampsia because they did not attend antenatal care due to the distance from their homes to health facilities.

> *“I don’t know much because I don’t go for antenatal care when I am pregnant” (Mother, 30-35 years)*.

Additionally, some participants attributed pre-eclampsia to a sedentary lifestyle:

> *“It also happens to people who sit a lot and do not do enough exercise”, (Mother, 30-35 years)*.

### Subtheme two: Signs and symptoms of pre-eclampsia

The study revealed that most mothers associated preeclampsia primarily with swollen feet, which was recognized as a common symptom during pregnancy. Additional frequently mentioned signs and symptoms included body weakness, poor vision, palpitations, and generalized headaches.

For example, one participant remarked:

> *“If someone has swollen feet, I think their pressure is also high. Sometimes, if the pressure is high, they also get a headache” (Mother, 20-24 years)*.

Another participant described a range of symptoms:

> *“Generalised body weakness, poor vision, a fast heartbeat, headache, and generalized body swelling” (Mother, 26-29 years)*.

Dizziness and excessive weight gain during pregnancy were also commonly cited as significant signs and symptoms of pre-eclampsia.

> *“Dizziness, leg swelling, increased heartbeats and being overweight” (Mother, 20-24 years)*.

### Theme two: Perceptions and stereotypes about pre-eclampsia

The study uncovered a range of perceptions and stereotypes about preeclampsia that significantly influenced health-seeking behaviour within the community. Many participants associated pre-eclampsia with witchcraft, while others referred to it as “Makiro,” a term used to describe swelling of the legs and other body parts after having sexual relations with multiple partners.

For instance, one participant remarked:

> *“People think it’s witchcraft, and others don’t know it completely” (Mother, 20-24 years)*.

Another participant explained:

> *“Those deep in the villages think it’s due to lack of exercise, witchcraft or usual pressure” (Mother, 30-35 years)*.

Additionally, a participant described the belief that pre-eclampsia was related to extramarital sexual activity:

> *“Makiro that a pregnant woman gets when her husband has sex with another woman” (Mother, 25-29 years)*.

Conversely, some mothers recognized that pre-eclampsia was more likely among women with twin pregnancies or multiple pregnancies. They noted that these pregnancies often start with leg swelling, which is often followed by elevated blood pressure:

> *“It relates with twin pregnancy, especially those with oedema or swollen legs” (Mother, 30-35 years)*.

### Theme three: Management of pre-eclampsia

Three sub-themes emerged under this theme, including response to pre-eclampsia, medicines used in the management of pre-eclampsia, and the application of these medicines in the community.

### Sub-theme one: Community response to pre-eclampsia

The community’s response to pre-eclampsia varied significantly. Some individuals sought assistance from village health teams and encouraged others to seek care at nearby health facilities if symptoms were identified.

For example:

> *“Mothers should be informed about how to manage the condition” (Mother, 25-29 years)*.

> *“Encourages her to go the hospital so as they see if it’s normal or not, specifically antenatal” (Mother, 20-24 years)*.

However, most of the participants preferred to consult traditional birth attendants and herbalists since they did not believe in the power of modern medicine:

> *“Yes, the herbalists as they believe modern medicine doesn’t work or go to diviners especially Local people” (Mother, 40-45 year)*.

> *“Village health workers can help them, together with traditional birth attendants and get some plant stems, (Mother, 30-35 years)*.

Additionally, some mothers mentioned using a drug called “Fukanga” sold by Chinese vendors in their village. Despite its perceived effectiveness, they noted that it was expensive:

> *“Fukanga is the best drug sold by Chinese but it’s very expensive,” (Mother, 40-45 years)*.

### Subtheme two: Medicines used in the management of pre-eclampsia

The study revealed that mothers in the community used various medicines for managing preeclampsia. These included traditional or unconventional remedies such as soap and onions, as well as plant-based solutions like *Bidens pilosa*, mumbwa, and roots mixed with avocado leaves.

Participants described their practices as follows:

> *“Me I normally use soap together with onions, I mix them, put them in water and drink the water twice a day, “(Mother, 30-35 years)*.

> *“Plants like Bidens pilosa have done me wonders, I even don’t waste my time going to the hospital, “(Mother, 30-35 years)*.

> *“Roots of plants that are pounded with dry avocado and Mumbwa work well, especially in the last trimester” (Mother, 36-39 years)*.

### Sub-theme three: Application of medicines used for the treatment of pre-eclampsia

The study revealed various methods employed by mothers in the community for the treatment of preeclampsia. Some preferred oral consumption of remedies, while others used topical applications on the stomach, and a few relied on practices such as sitting in the preparations, aimed at easing delivery and reducing the swellings.

Participants described their methods as follows:

> *“Just massaging it at least twice a day on both abdomen and feet”, (Mother, 30-35 years)*.

> *“Sitting in them to break bones and take orally”, (Mother, 30-35 years, Nkoma Village). “Some herbs and soil are added to water for the mother to bath”, (Mother, 40-45years)*.

Regardless of the method of application, timing was also another factor of factor. Most of the remedies were used either very early in the morning or late in the evening, depending on the type and mode of application.

> *“Take very early in the morning or cook and swallow. Just rubbing it without showering. The one for showering, you should use it very early in the morning. The one for smearing on the legs use it in the evening”, (Mother, 30-35 years)*.

### Theme four: Prevention of pre-eclampsia

The community employed various strategies to prevent pre-eclampsia, including lifestyle modifications and health practices. Key preventive measures included managing stress, maintaining a healthy diet, engaging in regular exercise, and advocating antenatal care. For those unable to access health facilities, consulting traditional birth attendants was also mentioned.

Participants emphasized:

> *“Avoid stress and fatty foods, do exercises, monitor their BP and go to the hospital early” (Mother, 36-40 years)*.

> *“Encouraging pregnant women to attend antenatal care and also health workers sensitize the women” (Mother, 25-29 years)*.

> *“Starting taking folic acid, exercise to have good blood circulation, going for antenatal visits, have a balanced diet,”(Mother, 30-35 years)*.

> *“If they have any complications, go for check-ups, do not to go the TBAs because this can cause complications,” (Mother, 24-29 years)*.

## Discussion

This study explored community perceptions and management practices regarding pre-eclampsia among women of reproductive age in Mbale City in Eastern Uganda. Our findings revealed a general lack of accurate knowledge about pre-eclampsia. Many women associated pre-eclampsia with symptoms such as swollen feet, headaches and body weakness. While most participants recognized pre-eclampsia as a condition associated with high blood pressure, a specific local term was absent for pre-eclampsia or eclampsia within the predominantly Bagisu community.

This contrasts with anecdotal reports from midwives practicing in the same locality who claim pre-eclampsia is sometimes referred to as ‘amakiro’. The community’s response to pre-eclampsia varied, with some relying on village health teams and traditional birth attendants, while others resorted to unconventional remedies such as soap and onion mixtures, plant-based solutions, and specific herbal medicines. Our findings align with previous research indicating low community awareness of pre-eclampsia in many low-income settings. Similar to studies in other sub-Saharan countries, participants often attributed symptoms of pre-eclampsia to stress, multiple pregnancies and high blood pressure, reflecting both accurate and misconceived notions of the condition. Our study’s findings on misconceptions about pre-eclampsia’s causes and risk factors are consistent with other research in sub–Saharan Africa. Misconceptions such as attributing eclampsia to anaemia, large babies, poor nutrition, witchcraft, and marital stress reflect patterns observed in Nigeria and Mozambique. In Nigeria, mothers attributed marital conflict, abusive husbands and strained relationships as responsible for pre-eclampsia [7]. In Mozambique, mothers believed that pre-eclampsia is caused by stress, worry and mistreatment from in-laws [2] and, extreme suggestions such as snakes living inside the woman’s body were fronted as possible explanations. Witchcraft was mentioned as a possible cause in our study and the Mozambican one [2][8]. These misconceptions often lead to delays in seeking appropriate care and reliance on traditional ineffective remedies. Despite these prevailing misconceptions, some participants correctly associated pre-eclampsia with high blood pressure. The lack of a distinct local term for pre-eclampsia in our study aligns with findings from similar settings such as Nigeria and Pakistan. In rural Nigeria, among the Yoruba, pre-eclampsia was described using analogies to other similar medical conditions, such as the “epilepsy of pregnancy” referring to eclampsia, the worst complication spectrum of pre-eclampsia that manifests with convulsions similar to epilepsy [9]. This pattern suggests a broader issue in the effective communication and management of pre-eclampsia, as the absence of a local term may impede recognition and intervention. A local name attached to pre-eclampsia with an identifiable set of symptoms is desirable as this gives the condition a local identity and enhances the uptake of interventions to manage it.

There were several misconceptions regarding the remedies for preeclampsia. The local remedy most participants easily turned to was herbal remedies. Herbal remedies are common throughout sub-Saharan Africa as a first mode of treatment. The concern is these remedies provide false hope and may cause delays in seeking treatment. One study in Nigeria found that the use of herbs was associated with severe eclampsia and preeclampsia[4]. Mothers also suggested prayer as a potential remedy. While there are strong religious beliefs in sub-Saharan Africa, these may contribute to the delay in seeking care or the first delay[10]. The reliance on traditional remedies, such as herbal treatments and unconventional mixtures, mirrors observations in rural Kenya and other regions [7] [8].

The misconceptions and reliance on traditional medicine identified in our study highlight the urgent need for targeted educational interventions about pre-eclampsia. Health promotion strategies should focus on demystifying the condition, addressing prevalent myths, and enhancing accurate knowledge among community members. Improving awareness about pre-eclampsia and its risks can lead to increased utilization of antenatal care services and timely medical interventions. Community health workers, equipped with this knowledge, could play a pivotal role in bridging the gap between traditional practices and modern healthcare.

Respondents in our study were aware of the dangers of eclampsia and expressed fear of maternal death as the ultimate consequence of the condition. This awareness provides a window of opportunity for interventions to target the broader challenge of misconceptions. The Health Belief Model has been used to explain poor health care-seeking behaviours in Nigeria owing to misconceptions [10]. The perceived susceptibility to stillbirth and death among mothers with eclampsia, as reported by some participants in our study, and the acknowledgement by some that medical attention should be sought, could provide a valid basis for interventions [4]. Community health workers have shown sufficient knowledge and ability to identify women with pre-eclampsia and administer initial treatment [13]. Future programs will need to develop interventions that focus on demystifying the prevalent myths and promoting a more scientific understanding of the condition, which could eventually serve as cues for action [14].

Future research should focus on evaluating the effectiveness of community-based educational interventions in improving knowledge and management of pre-eclampsia. Longitudinal studies could explore the impact of enhanced antenatal care services on pre-eclampsia prevalence and outcomes in similar settings.

### Strengths and Limitations

A major strength of our study is its qualitative approach, which provided in-depth insights into community perceptions and management practices. However, the study also has limitations. Data were collected exclusively through in-depth interviews, which could have been complemented by focus group discussions for a richer understanding.

## Conclusions

Our study in rural Eastern Uganda highlights a critical need for enhanced awareness and education about pre-eclampsia. The absence of a clear local term and prevalent myths can negatively influence health-seeking behaviour and delay appropriate care. Addressing these gaps through culturally sensitive educational programs, improving access to antenatal care, and facilitating timely referrals are essential steps for the early detection and effective management of pre-eclampsia. Such measures are crucial for improving maternal and neonatal health outcomes in low-income settings.

## Data Availability

University Repository

## Acknowledgements

We extend our gratitude to Seed Global Health for organizing the community engagement outreach program that raised awareness about pre-eclampsia and provided a platform for conducting this study. We also acknowledge the local leadership of Mbale City for their invaluable support during this exercise. Additionally, we thank the Busitema University Students’ Association for its dedication to championing community engagement efforts.

## Consent for publication

Not applicable

## Availability of data and materials

The datasets used and/or analyzed during the current study are available from the corresponding author on reasonable request.

## Funding

There was no funding for this study.

## Notes

**Conflict of interest** The authors declare that they have no competing or conflicts of interest.

### Competing Interest Statement

The authors have declared no competing interest.

### Funding Statement

The author(s) received no specific funding for this work.

### Author Declarations

Ethical approval was obtained from the Research Ethics Committees (REC) of Mbale Regional Referral Hospital (approval number MRRH-2023-300.

## References

[1] W. Wang et al., “Epidemiological trends of maternal hypertensive disorders of pregnancy at the global, regional, and national levels: a population_Jbased study,” BMC Pregnancy Childbirth, vol. 21, no. 1, pp. 1–10, 2021, doi: 10.1186/s12884-021-03809-2.

[2] World Health Organisation (WHO), “Analytical Fact Sheet Maternal mortality: The urgency of a systemic and multisectoral approach in mitigating maternal deaths in Africa Rationale,” Anal. Fact Sheet, no. March, pp. 1–11, 2023.

[3] S. Awor et al., “Prediction of pre-eclampsia at St. Mary’s hospital lacor, a low-resource setting in northern Uganda, a prospective cohort study,” BMC Pregnancy Childbirth, vol. 23, no. 1, pp. 1–10, 2023, doi: 10.1186/s12884-023-05420-z.

[4] C. Ndwiga, G. Odwe, S. Pooja, O. Ogutu, A. Osoti, and C. E. Warren, “Clinical presentation and outcomes of preeclampsia and eclampsia at a national hospital, Kenya: A retrospective cohort study,” PLoS One, vol. 15, no. 6 June, Jun. 2020, doi: 10.1371/JOURNAL.PONE.0233323.

[5] B. Haddad and B. M. Sibai, “Expectant Management in Pregnancies with Severe Pre-eclampsia,” Semin. Perinatol., vol. 33, no. 3, pp. 143–151, Jun. 2009, doi: 10.1053/j.semperi.2009.02.002.

[6] L. Duley, “The Global Impact of Pre-eclampsia and Eclampsia,” Semin. Perinatol., vol. 33, no. 3, pp. 130–137, Jun. 2009, doi: 10.1053/J.SEMPERI.2009.02.010.

[7] A. Degu Ayele and Z. A. Tilahun, “Magnitude of Preeclampsia and Associated Factors Among Women Attending Delivery Service in Debre Tabor Specialized Hospital,” 2022, doi: 10.4314/ejhs.v32i2.8.

[8] M.W. O’Hara and A. M. Swain, “Rates and risk of postpartum depression - A meta-analysis,” Int. Rev. Psychiatry, vol. 8, no. 1, pp. 37–54, 1996, doi: 10.3109/09540269609037816.

[9] J. Quist-Nelson et al., “Early magnesium discontinuation postpartum and eclampsia risk: A systematic review and meta-analysis,” Pregnancy Hypertens., vol. 37, Sep. 2024, doi: 10.1016/j.preghy.2024.101141.

[10] C. V. Ananth, D. A. Savitz, and W. A. Bowes, “Hypertensive disorders of pregnancy and stillbirth in North Carolina, 1988 to 1991,” Acta Obstet. Gynecol. Scand., vol. 74, no. 10, pp. 788–793, 1995, doi: 10.3109/00016349509021198.

